# Prevalence of the Cefazolin Inoculum Effect (CzIE) in Nasal Colonizing Methicillin-Susceptible *Staphylococcus aureus* in Patients from Intensive Care Units in Colombia and Use of a Modified Rapid Nitrocefin Test for Detection

**DOI:** 10.1101/2024.07.11.24309236

**Authors:** Lina P. Carvajal, Sandra Rincon, Sara I. Gomez-Villegas, Juan M. Matiz-González, Karen Ordoñez, Alejandra Santamaria, Leonardo Ospina-Navarro, Jaime Beltran, Fredy Guevara, Yardany R. Mendez, Soraya Salcedo, Alexandra Porras, Albert Valencia-Moreno, Haley Grennia, Alexander Deyanov, Rodrigo Baptista, Vincent H. Tam, Diana Panesso, Truc T. Tran, William R. Miller, Cesar A. Arias, Jinnethe Reyes

**Affiliations:** Molecular Genetics and Antimicrobial Resistance Unit, Universidad El Bosque, Bogota, Colombia; Brigham and Women’s Hospital, Harvard Medical School, Boston, MA, USA; Department of Infectious Diseases, ESE Hospital Universitario, San Jorge de Pereira, Pereira, Colombia; Clinica Colsubsidio Calle 100, Bogota, Colombia; Servicio de Infectología, Fundación Santafe de Bogota, Bogota, Colombia; Clinica Reina Sofia, Colsanitas, Bogota, Colombia; Grupo de Investigacion en Epidemiologia Clinica de Colombia (GRECO), Universidad Pedagogica y Tecnologica de Colombia, Tunja, Colombia; Hospital Regional de Duitama, Duitama, Colombia; Organizacion Clinica General del Norte, Barranquilla, Colombia; Los Cobos Medical Center, Bogota, Colombia; Center for Infectious Disease, Houston Methodist Research Institute, Houston, TX USA; Division of Infectious Diseases and Department of Medicine, Houston Methodist Hospital, Houston, TX USA 77030; Department of Medicine, Weill Cornell Medical College, New York, NY.; Departament of Pharmacy Practice and Translational Research, University of Houston, Houston, Texas, United States

## Abstract

The cefazolin inoculum effect (CzIE) has been associated with poor clinical outcomes in patients with MSSA infections. We aimed to investigate the point prevalence of the CzIE among nasal colonizing MSSA isolates from ICU patients in a multicenter study in Colombia (2019-2023). Patients underwent nasal swabs to assess for *S. aureus* colonization on admission to the ICU and some individuals had follow-up swabs. We performed cefazolin MIC by broth-microdilution using standard and high-inoculum and developed a modified nitrocefin-based rapid test to detect the CzIE. Whole genome sequencing was carried out to characterize BlaZ types and allotypes, phylogenomics and Agr-typing. All swabs were subjected to 16S-rRNA metabarcoding sequencing to evaluate microbiome characteristics associated with the CzIE. A total of 352 patients were included; 46/352 (13%) patients were colonized with *S. aureus*; 22% (10/46) and 78% (36/46) with MRSA and MSSA, respectively. Among 36 patients that contributed with 43 MSSA colonizing isolates, 21/36 (58%) had MSSA exhibiting the CzIE. BlaZ type A and BlaZ-2 were the predominant type and allotype in 56% and 52%, respectively. MSSA belonging to CC30 were highly associated with the CzIE and SNP analyses supported transmission of MSSA exhibiting the CzIE among some patients of the same unit. The modified nitrocefin rapid test had 100%, 94.4% and 97.7% sensitivity, specificity and accuracy, respectively. We found a high prevalence point prevalence of the CzIE in MSSA colonizing the nares of critically-ill patients in Colombia. A modified rapid test was highly accurate in detecting the CzIE in this patient population.

## INTRODUCTION

*Staphylococcus aureus* is a human pathogen and a leading cause of community and healthcare-associated infections (HAI). *Staphylococcus* species (*S. aureus* and coagulase negative staphylococci [CoNS]) reside in similar ecological niches in the human body, nasal cavity, nasopharynx, skin, and mucous membranes.^1–4^ It is estimated that about 20% (range 12–30%) of the healthy human population is permanently colonized with *S. aureus*, 30% of which are intermittent carriers (range 16-70%) and 20% are regarded as non-colonized.^1–4^ The current evidence suggests that there is variability in colonization rates due to geographical location and characteristics of the tested populations.^1^

Colonization by *S. aureus* is a well-recognized risk factor for the development of subsequent infections, in particular, bacteremia and hospital-associated pneumonia.^5–7^ At risk patients include those admitted to the ICU, representing a vulnerable group of individuals with major organic dysfunctions, comorbidities and receiving prolonged courses of broad-spectrum antibiotics. For example, nasal colonization with methicillin-resistant *S. aureus* (MRSA) is a known risk factor for developing ventilator-associated pneumonia due to these organisms.^8^ Furthermore, screening for multidrug-resistant organisms (MDRO) is often performed in these vulnerable patient populations to guide isolation policies and decolonization strategies.^9^

Information on the rates of *S. aureus* colonization in patients in the ICU setting in developing countries is limited. Olarte et al., reported that in a population of 708 patients from one ICU in Bogota, Colombia, the rate of *S. aureus* colonization at admission was 25.8% (7.2% for MRSA and 18.5% for MSSA).^10^ In a prospective Latin-American study that included a cohort of 675 patients enrolled between 2011 and 2014 in hospitals across nine countries of the region, MSSA were responsible for a substantial number (53.7%) of cases of bacteremia.^11^ Aggregated data from the Colombian National Health Institute indicates that *S. aureus* is the fourth and third most common pathogens causing hospital-associated and ICU-related infections, respectively.^12^ MSSA accounted for 66% of the infections in the Colombian ICUs.^12^

β-lactams are the first line therapy for MSSA infections and, with cefazolin frequently being the “go-to” choice in deep-seated infections.^13^ This preference is largely due to its better side effect profile and tolerability, longer half-life, improved adherence, and low costs, and comparable outcomes.^14–16^ In addition, cefazolin is also the first-choice option for surgical prophylaxis in a variety of surgical interventions including cardiac to orthopedic surgery.^17–18^ However, a concern with using cefazolin is the presence of the inoculum effect (CzIE). Indeed, the CzIE has been associated with therapeutic failures in patients with MSSA bloodstream infections,^19,20^ and with the progression to chronic osteomyelitis in children.^21^ In Latin-America, a high prevalence of the CzIE among clinical MSSA isolates recovered from invasive infections has been documented.^22,23^ Studies have documented rates of the CzIE in Colombia ranging from 34% to 63% in bloodstream and osteomyelitis infections, respectively.^23^ A more recent study focusing on bloodstream isolates from reported a point prevalence of 40% for the CzIE.^22^

In this work, we postulated that the prevalence of the CzIE in colonizing MSSA isolates recovered from patients in the ICU would be higher compared to other patient populations, likely due to increased use of antimicrobials (particularly β-lactams) in these patients. Thus, we conducted a multicenter study to investigate the point prevalence of the CzIE among nasal colonizing MSSA isolates from ICU patients in four different cities in Colombia. Additionally, we tested the ability of a modified colorimetric test to identify the CzIE in MSSA recovered from ICU patients.

## MATERIALS AND METHODS

### Patient enrollment and nasal swab sampling

We performed a prospective observational cohort study that included 352 patients admitted to six adult ICUs in six high complexity-hospitals in Colombia, enrolled between October 2019 to August 2023, (Universidad El Bosque IRB protocol 476-2018). The Human Subjects Review Committee of the participant institutions approved the study. Patients or their authorized representative provided signed consent for participation in the study. Inclusion criteria encompassed adult patients (>18 years old), admitted at ICU, and provided signed consent. Exclusion criteria encompassed patients from whom samples were not collected upon ICU admission (or those from whom samples were obtained >24 hours after admission), and patients diagnosed with COVID-19. Participating centers (affiliated with medical schools and private hospitals) were in Bogota, (3 hospitals; hospitals C, D, E), Pereira (1 hospital; hospital B), Barranquilla (1 hospital; hospital A), and Duitama (1 hospital; hospital G). Two nasal swabs were obtained from the anterior nares (one for microbiological screening of multidrug-resistant pathogens (MDRO) and one for microbiome analysis) at 24 hours of admission to the ICU. Samples were taken by trained nurses and physicians, who were members of the participating centers. Nasal samples were obtained from both nostrils and taken using swabs placed immediately in AIMES transport media (to collect and preserve the nasal samples for microbiological screening) and DNA/RNA Shield to stabilize nucleic acids for microbiome analyses. These samples were kept at 4°C until they were shipped to the reference laboratory at Universidad El Bosque. MDRO pathogens included Carbapenem-Resistant (CR) *Klebsiella pneumoniae*, Enterobacterales, *Pseudomonas* and *Acinetobacter*. Methicillin-Resistant *Staphylococcus aureus*, and Vancomycin-Resistant *Enterococcus* using chromogenic agar media (CHROMagar-Orientation). Subsequently, a follow-up nasal swab was planned to be collected from the patients before ICU discharge or hospital transfer (day 1 to day 24), unless the patient died during the ICU stay before a sample could be taken or if there were personnel were unable to collect the sample. However, only 175 out of 352 patients had follow-up nasal swabs.

### Microbiological screening

Nasal swabs were inoculated onto BHI agar and incubated overnight. Then, using a 10 μL calibrated loop, a loopful of bacterial culture was transferred to 5 mL of BHI broth that was incubated for 2 hours at 37°C. Serial dilutions of the BHI broth were performed on saline solution. 500 μL of a 10^-5^ dilution was spread onto Chromogenic agar media (CHROMagar-Orientation) using a sterile glass spreader and incubated at 37°C for 18 hours. Colonies compatible with *S. aureus* were recovered and sub-cultured on BHI agar and identified at genus-species level using a multiplex PCR, which also confirmed the presence of the *mecA* gene.^24^ Further, oxacillin susceptibility testing was performed using the agar dilution method following CLSI guidelines.^25^

### Cefazolin high inoculum effect determination

Cefazolin susceptibility testing was performed by broth microdilution using a standard inoculum of 5x10^5^ CFU/mL following CLSI guidelines.^26^ Additionally, cefazolin MICs were determined using a high bacterial inoculum (5x10^7^ CFU/mL), as previously described,^22,27^ in three biological replicates. Cefazolin concentrations were from 0.03 to 64 mg/L. Reading of the MIC values at high inoculum and interpretation of the results was carried-out by three independent investigators. The CzIE was defined as MIC of ≤8 mg/L at standard inoculum and ≥16 mg/L at high bacterial inoculum.

### Modification of the rapid test protocol to identify MSSA with CzIE

We used a previously published^28^ rapid nitrocefin test to identify isolates exhibiting the CzIE with a modification using ampicillin disks (10 μg; Oxoid) instead of ampicillin solution.^29^ Briefly, 1 mL of BHI broth (Oxoid) and one disk of ampicillin (10 μg) were combined in a 2 mL tube, vigorously mixed for at least 30 seconds and incubated for 10 min at room temperature. The assay required that the ampicillin disk was completely submerged into the broth. Next, the solution was mixed vigorously for 30 seconds, and the disk was then removed from the broth using sterile loop or tweezer, leaving 1 mL solution of BHI broth containing ampicillin ready to use. Colonies from a pure and fresh culture (less than 72 hours) with similar morphological features were selected on BHI agar using a 1-μL calibrated sterile loop. Then, the content of 1 μL loop full of bacterial colonies was resuspended in 1 mL of ampicillin solution followed by vigorous vortexing for 2 min and incubated for 10 min at 36±1°C. Subsequently, the bacterial suspension was vortexed again for 2 min. Then, a second incubation of 10 min at 35 ±1°C followed by 2 minutes of vigorous vortexing was performed. Next, the solution was centrifuged at 5,000 rpm for 10 min at room temperature. A volume of 25 μL of the supernatant was transferred into a 0.2-mL tube containing 25 μL of freshly prepared nitrocefin solution (stock concentration 400 μM prepared in phosphate-buffered saline [PBS] at 0.1 M, pH 7.0, as previously reported).^28,29^ This process ensured that the bacterial cell pellet at the bottom of the tube was not disturbed. Subsequently, the nitrocefin tubes were incubated at room temperature (15 to 25°C, mean of 22°C) and protected from light, followed by visual inspection to monitor color change at 30 min, 1 h, and 2 h. The results were interpreted under the incident light and compared with controls (see below). A positive result was any observed change in color (from yellow to red) between 30 min to 2 h (See Supplemental Material) (**Fig.1**). Quality control strains ATCC *S. aureus* 29213 (lacking the CzIE and with a negative rapid test) and TX0117 (exhibiting the CzIE and with a positive modified rapid test) were included as control strains, as previously described.^22,28,29^ Performance metrics (sensitivity, specificity, and accuracy) of the modified rapid test were compared to MIC determination at high inoculum (gold standard) using MedCalc software.

**Fig. 1.**
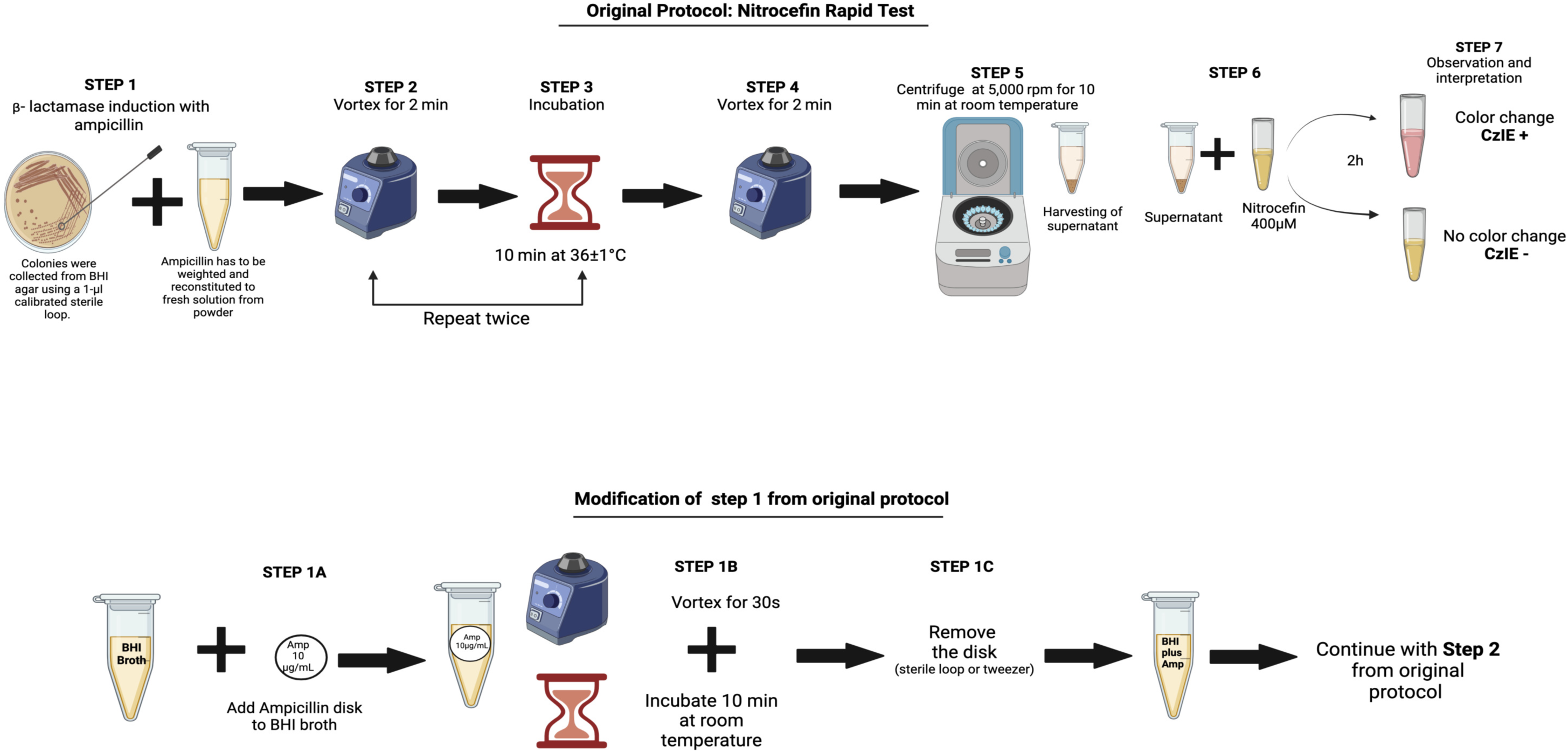
Modified protocol of the nitrocefin rapid test to identify MSSA with the CzIE.

### Whole genome sequencing, assembly, and genomic analysis

Bacterial DNA extraction was carried out on overnight BHI cultures after lysostaphin treatment using a Qiagen Kit. Library preparation was performed using Illumina prepkit, and sequencing was performed on the Illumina platform NextSeq 2000. The raw reads obtained from sequencing by Illumina MiSeq were subjected to quality control by FastQC and Trimmomatic.^30,31^ Subsequently, the reads were assembled with SPAdes,^32^ annotation was performed using PROKKA^33^ and the identification of species was verified employing StrainSeeker, KmerFinder and MLST. Sequence Type (ST) and Clonal Complex identification, as well as submission and curation of new STs, was performed on the PubMLST platform.

Identification of the *blaZ* gene was performed by BLASTn using the sequence from the strain *S. aureus* ATCC 29213 extracted with SeqKit^34^ and translated using MEGAX.^35^ BlaZ typing was carried out taking into account the amino acids at positions 128 and 216, corresponding to positions 119 and 207 including the signal peptide, as we previously described.^22^ Finally, the assignment of BlaZ allotypes (unique amino acid sequences in the analyzed genomes) was performed as previously described using an identity matrix for protein sequence comparison elaborated with the previously described 29 allotypes. ^22^

The presence of virulence determinants as accessory gene regulator (Agr) was searched using *in silico* PCR,^36^ and BLASTn based on the sequence of the four reported *agr* loci (I-IV). Additionally, the Panton-Valentine Leukocidin (PVL) encoding genes (*lukS* and *lukF*) were searched by *in silico* PCR,^37^ and BLASTn using the coding sequences of *lukS* (ABD22441.1) and *lukF* (ABD21112.1) from *Staphylococcus aureus* subsp. *aureus* USA300_FPR3757.

### Phylogenomic analysis

Phylogenetic inference of the nasal MSSA genomes was performed using the core genome (orthologous genes present in at least 95% of the genomes) obtained with Roary,^38^ using a nasal colonizer *S. argenteus* genome as the outgroup recovered in this work. Orthogroups were individually aligned with PRANK,^39^ and then concatenated to obtain a consensus matrix. The maximum likelihood (ML) tree was built with the consensus matrix in RAxML^40^ using the GTR evolutionary model and a branch support analysis of 1,000 bootstrap replicates. The resulting ML phylogenetic tree was plotted using iTOL.^41^

### Library preparation and 16S sequencing

Among the 36 patients colonized with MSSA, twenty-three nasal swabs collected at admission to ICU (10 patients colonized by MSSA exhibiting the CzIE and 13 by MSSA lacking the CzIE) were available for microbiome analysis. Total DNA was obtained using DNA easy Blood and Tissue kit (QIAGEN). DNA samples were subjected to bacterial diversity characterization by amplicon sequencing of the hypervariable region V4 of the 16S rRNA.^42^ For the sequencing process, microbial amplicon libraries were built using barcode primers 515-F (5ʹ-GTG CCA GCM GCC GCG GTA A-3ʹ) and 806-R (5ʹ-GGA CTA CHV GGG TWT CTA AT-3ʹ).^43^ After sequencing, forward and reverse raw reads were demultiplexed and merged after primer removal using bcl2fastqc software (Illumina). The Quantitative Insights into Microbial Ecology (QIIME2) program was used to extract barcodes and primers from the resulting paired-end sequences.^44^ Then, DADA2 (QIIME2 plugin) was used to remove chimera reads and evaluate the quality of the resulting reads with a Q-Score plot to select the minimum length with an average quality ≥ 30. In addition, DADA2 was also used to infer amplicon sequence variants (ASV) by clustering all identical sequenced reads, and taxonomic assignment of ASVs was performed using the Silva v132 database.^45^ Finally, a read abundance matrix of the microbial groups identified for each sample, at the phylum or genus level, was generated.

The resulting matrix was used for the calculation of a Bray-Curtis dissimilarity matrix that was utilized for PCoA analysis using the ordinate and plot_ordination functions. Differences between groups were evaluated by PERMANOVA and ANOSIM analysis using adonis2 and anosim functions, respectively. Furthermore, microbial α-diversity metrics (Shannon and Simpson indexes, and Richness) were calculated with the function estimate_richness, using the abundance matrix rarefied per sample to 2000 reads through rarefy_even_depth function. All functions are contained in the Phyloseq and Vegan R packages.^46,47^ Taxonomic relative abundance matrices were employed to describe general microbiome composition (all phyla/genera are showed, except taxa with a relative abundance < 0.5%) in CzIE-positive MSSA and CzIE-negative MSSA nasal microbiome groups. In addition, a differential abundance taxa analysis between groups was performed via run_lefSe function of the Microbiome Marker R package, using default options (p-value < 0.05 and an LDA > 2 or LDA < −2).^48^ All graphs and statistical comparisons (Mann-Whitney tests) were done using the Graph Pad Prism software v8.0.1.

### Core SNPs Analysis

The core genome SNPs of the 43 MSSA nasal isolates was built with kSNP4 (https://sourceforge.net/projects/ksnp/), using a kmer size set to 19, the optimum size estimated by the kSNP4 program Kchooser.^49^ Pairwise core SNP distance matrix was constructed with the core SNPs using the SNP-dist script (https://github.com/tseemann/snp-dists).

### Data availability

The genomes of the Methicillin-Susceptible *Staphylococcus aureus* isolates, and the nasal microbiome sequencing were submitted to the NCBI GenBank database under the BioProject numbers PRJNA1107600 and PRJNA1108236, respectively.

## RESULTS

### Colonizing MSSA isolates from ICU patients showed a high prevalence of the CzIE

A total of 352 patients were included in the study from 2019 to 2023. The patients were enrolled in six ICUs located in four different cities of Colombia (Barranquilla, Pereira, Bogota and Duitama). Nasal swabs were obtained from all the 352 patients on admission at ICU. Subsequent nasal swabs were collected in 175 patients at follow-up (see Materials and Methods). 46/352 (13%) patients included in the study were colonized by *S. aureus*. Among those with *S. aureus* colonization, 22% (10/46) and 78% (36/46) were colonized by MRSA and MSSA, respectively (**Table 1 and Fig. 2**).

**Fig. 2.**
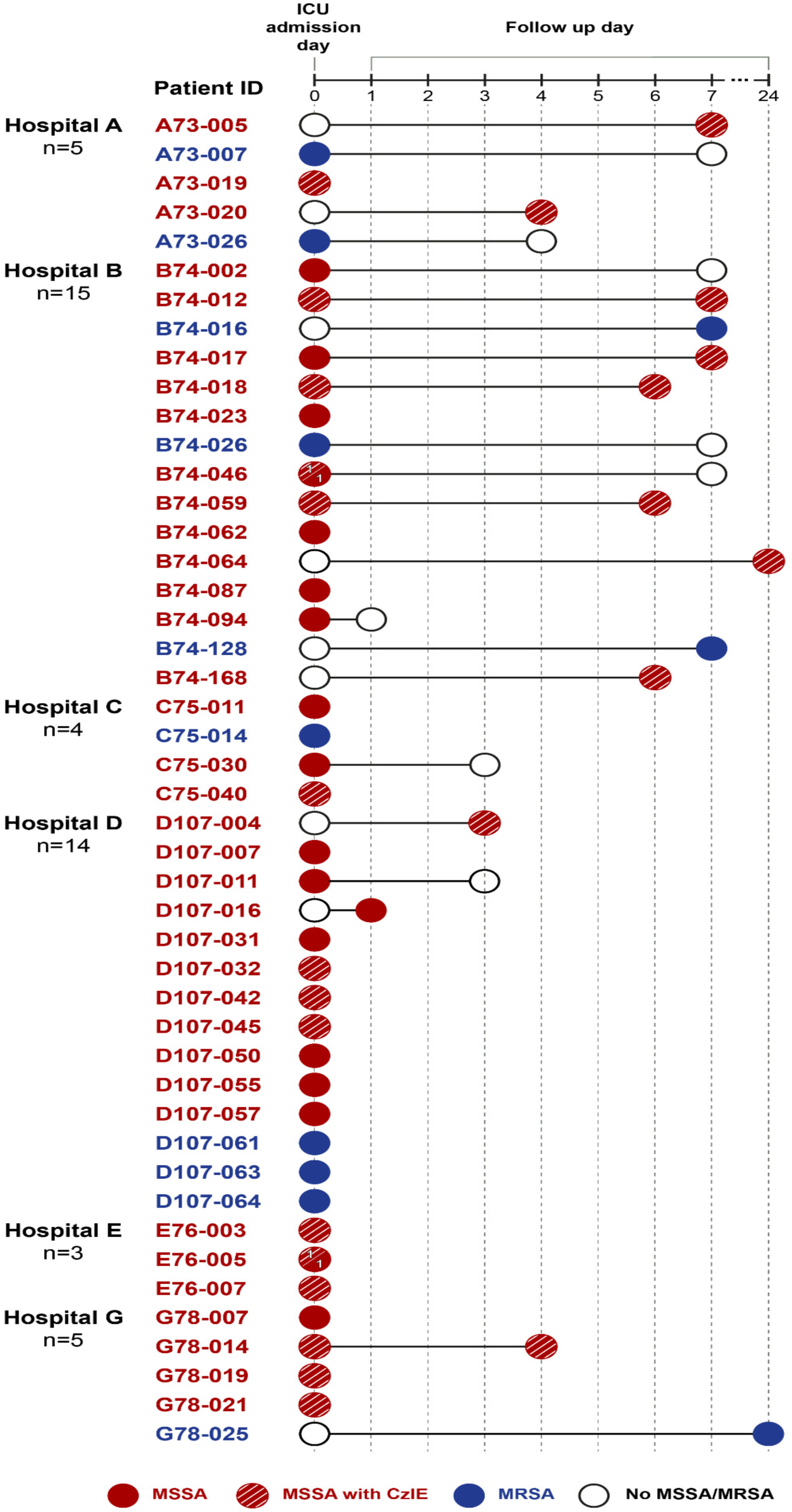
Timeline of sampling and isolate recovery in ICU patients colonized by *S. aureus* (MSSA or MRSA). Sampling is denoted by circles; filled circles represent nasal swabs positive for MSSA (red) or MRSA (blue); empty circles indicate samples negative for *S. aureus.* Circles with lines across indicate isolates positive for the CzIE. Patient B74-046 and E76-005 had two isolates recovered in the same swab, one displaying the CzIE and the other lacking the CzIE.

**Table 1.**
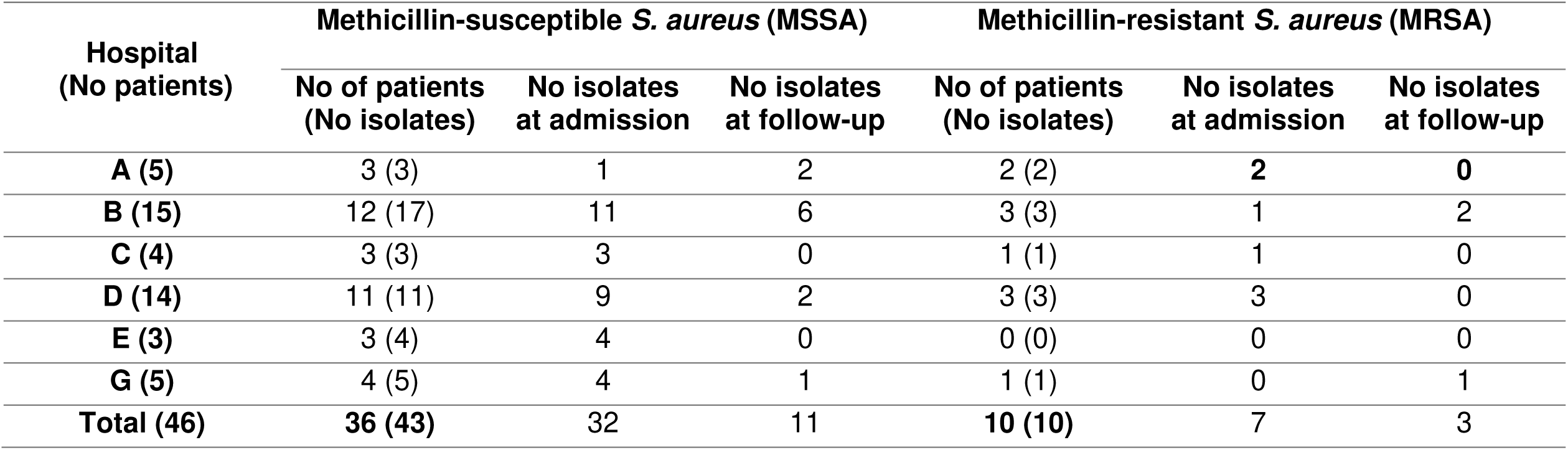
Distribution of *S. aureus* isolates recovered from 46 patients on admission and follow-up by hospital.

Among the 46 patients colonized with *S. aureus* who had a nasal swab taken at admission, 22 (48%) had an additional nasal swab taken in the follow-up period of time which varied from day 1 to 24 after admission (**Fig. 2**). A total of 37/46 patients (70%) were colonized by *S. aureus* in the first nasal swab upon admission to the ICU (30 and 7 with MSSA and MRSA, respectively) (**Fig. 2**). Among the 22 patients who underwent a follow up nasal swab, 9 were found to be colonized by *S. aureus* only in the second swab on days 1 to 24 (colonization was absent upon admission to the ICU), suggesting a potential acquisition during the hospital stay or lack of detection in the first swab (**Fig. 2**). Among the 22 patients, 13 were positive in the first swab and 8 negative in the second swab. Of note, 5 patients were colonized on admission and remained colonized on the second swab taken in the follow up period (only with MSSA) (**Fig. 2**). A total of 53 *Staphylococcus aureus* isolates were found in 46 patients (**Fig. 2**); 39 isolates (73%) recovered in nasal swabs taken on admission, (7 MRSA and 32 MSSA) and 14 (27%) isolates (3 MRSA and 11 MSSA) on follow up swabs (**Table 1 and Fig. 2**).

Among 36 patients that contributed 43 MSSA colonizing isolates, 21/36 (58%) had nasal colonization with MSSA exhibiting the CzIE (**Table 2 and Fig. 2**). Of these, 11 patients were colonized on admission to the ICU, 6 patients during the follow-up period only (range of 3 to 24 days) and 4 patients were colonized at admission and remained colonized during the follow-up (range 4 to 7 days) (**Fig. 2**). The CzIE was identified in 25/43 (58%) of nasal colonizing MSSA (geometric mean of cefazolin MIC of 41.07 mg/L, range 16-128 mg/L).

**Table 2.** Distribution of the frequency of the CzIE in colonizing MSSA recovered from ICU patients in 6 Colombian hospitals by gold standard testing.

| **Hospital** | **No of MSSA isolates** | **No of isolates (%) with CzIE** | **No of isolates (%) without CzIE** |
| --- | --- | --- | --- |
| **A** | 3 | 3 (100) | 0 (0) |
| **B** | 17 | 10 (59) | 7 (41) |
| **C** | 3 | 1 (33) | 2 (67) |
| **D** | 11 | 4 (36) | 7 (64) |
| **E** | 4 | 3 (75) | 1 (25) |
| **G** | 5 | 4 (80) | 1 (20) |
| **Total** | **43** | **25 (58)** | **18 (42)** |

### A modified rapid nitrocefin test identified the CzIE in nasal colonizing MSSA isolates with high accuracy

Due to logistic issues to obtain ampicillin powder for the rapid nitrocefin test previously published, we opted to modify the test by using ampicillin discs (10 μg) for β-lactamase (BlaZ) induction, since such disks are widely available in developing countries (See Supplemental Material).

Compared to the gold standard (MICs using high inoculum by broth microdilution), the modified rapid test identified colonizing MSSA isolates with the CzIE with a sensitivity of 100%, specificity of 94.4% and an overall accuracy of 97.7%. Table S1 shows that the test showed 100% sensitivity with the three common types of class A β-lactamases (Type A, B and C). The specificity was lower (87.5%) in isolates harboring type B β-lactamase. All colonizing MSSA isolates without *blaZ* gene (CzIE negative), were also correctly identified as negative with the modified CzIE test (**Table S1**).

### Genomic characteristics of nasal colonizing MSSA isolates from ICU

Whole genome sequencing of the 43 MSSA isolates indicated that 91% (39/43) of the nasal colonizing MSSA isolates carried the *blaZ* gene (**Fig. 3**). Among them, BlaZ type A and type C were identified in 41% (n=16/39), and 36% (n=14/39) of isolates, respectively. BlaZ type B was the less common type, detected in 23% (n=9/39) of MSSA carrying *blaZ*.

**Fig. 3.**
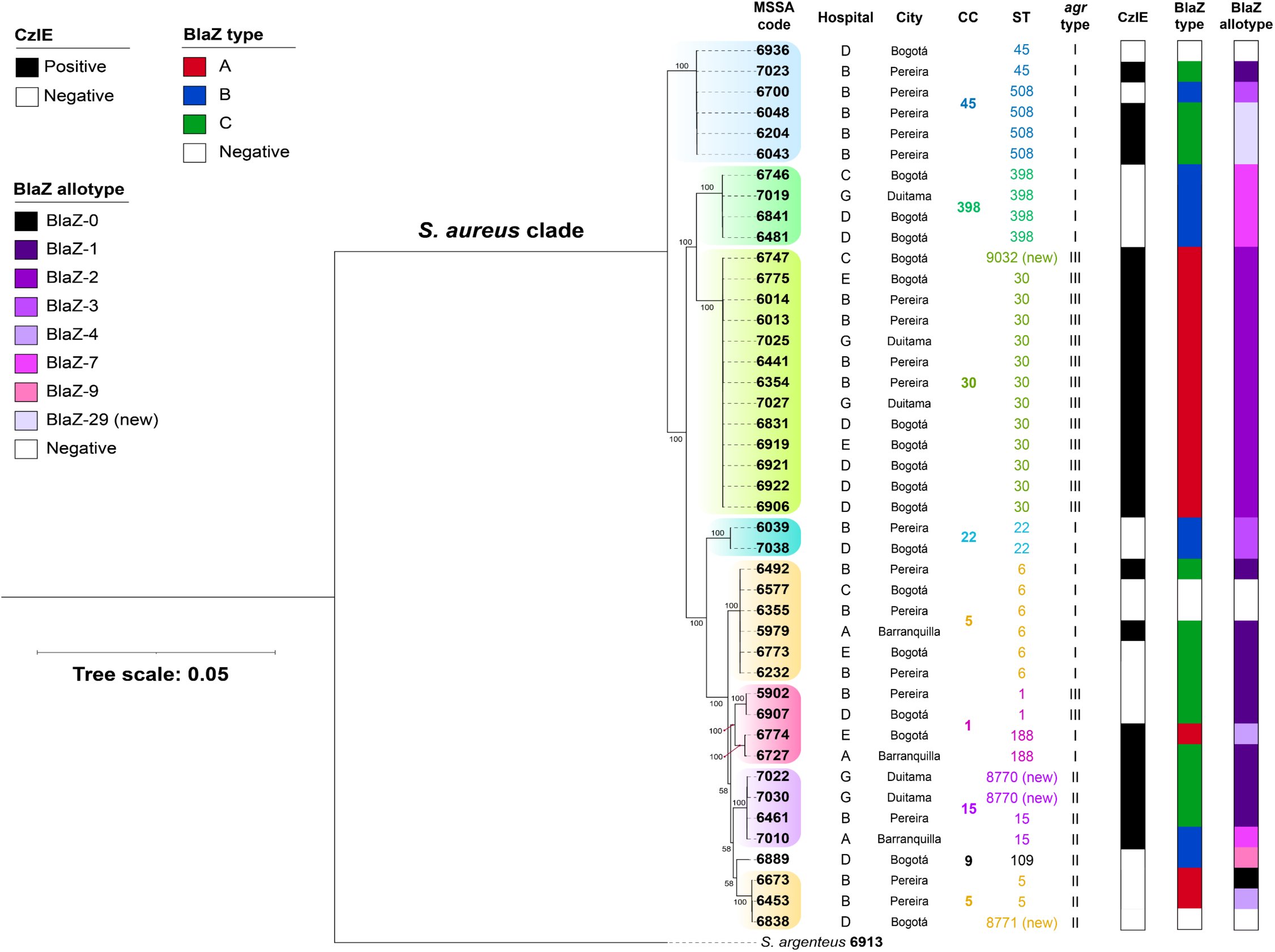
Phylogenetic tree of colonizing methicillin-susceptible *S. aureus* (MSSA) isolates recovered from ICU patients in Colombian hospitals. Maximum likelihood phylogenetic tree from the core genome (1939 genes) of 43 MSSA and one *S. argenteus* isolate (outgroup). Support bootstrap values are showed for each clade. Colored shadows over the tree branches show the clonal complexes (CC) within the sample, followed by Sequence Type (ST) and *agr* type. The CzHI was determined by gold standard (CFZ MIC)

When analyzing 25 nasal colonizing MSSA isolates that exhibited the CzIE, BlaZ type A was the predominant type in 56% (14/25 isolates), followed by BlaZ type C in 40% (n=10/25). BlaZ-2 was the predominant allotype in 52% (n=13/25). A novel allotype BlaZ-29 (derived from type C BlaZ) was detected in three isolates displaying the CzIE (**Fig. 3**).

Among 18 MSSA lacking the CzIE, 14 isolates harbored the *blaZ* gene and type B was the most predominant BlaZ type identified in 57% (8 out of 14) of these genomes. BlaZ-1 (from type C) and BlaZ-7 (from type B) were the most common allotypes detected in MSSA lacking the CzIE (57%, 4 isolates of each allotype) (**Fig. 3**).

We identified a high diversity of clonal lineages among 43 nasal colonizing MSSA isolates from ICU patients, represented by eight major clades related to the clonal complexes identified (CC1, CC5, CC9, CC15, CC22, CC30, CC45 and CC398) (**Fig. 3**). The MSSA that exhibited the CzIE (n = 25) clustered into five clonal groups, CC1 (n = 2), CC5 (n = 2), CC15 (n = 4), CC30 (n = 13), and CC45 (n = 4).

Notably, CC30 was the predominant lineage identified among MSSA displaying the CzIE (52%, 13 out of 25 isolates), with most of the isolates belonged to ST30 (12 out 13 CC30 isolates). Of note, all isolates displaying the CzIE and carrying Agr-III, belonged to CC30 and harbored BlaZ type A and allotype BlaZ-2. MSSA lacking the CzIE clustered into six clonal complexes, CC1 (n = 2), CC5 (n = 7), CC9 (n = 1), CC22 (n = 2), CC45 (n = 2), and CC398 (n = 4); CC5 was the most common genetic background, identified in 39% (7 out of 18 of the MSSA without the CzIE), followed by CC398 found in 22% (n=4 isolates). Further, we found that CC9, CC22, and CC398 were lineages exclusively detected in MSSA isolates lacking the CzIE (**Fig. 3**).

Phylogenomic analyses indicated that the MSSA population was heterogeneous with the absence of geographical or hospital associations with particular clades, except for a clade belonging to CC45 where five out of six MSSA were recovered from the same hospital in the city of Pereira (**Fig. 3**). The most common clonal cluster (CC) associated with the CzIE was CC30 (n=13/43). A minor clustering of isolates exhibiting the CzIE (n = 4) was observed in CC15.

Using the genomic data we also analyzed MSSA isolates recovered from the same patient to evaluate genetic relatedness between these isolates and if subsequent isolates acquired or lost the CzIE. A total of 5 patients had MSSA obtained from samples taken on day of admission as well as in the follow up time (4 to 7 days). Whereas two patients had two MSSA isolates recovered only in the day of admission (**Fig. 4 and Table S2**). Among the isolates exhibiting the CzIE and recovered from samples taken at different time points, 3 pairs of isolates (6013/6014, 6048/6204, and 7022/7030) (**Fig. 4 and Table S2**) showed high genomic relatedness with <20 SNPs difference, suggesting the same MSSA strain. In contrast, isolates 6039/6043, and 6441/6461 recovered from two patients at different time points, displayed distinct genomic, phenotypic, and phylogenetic features suggesting that the patients were colonized with different strains. Similarly, MSSA recovered from the same nasal sample on admission in two patients (isolates 6354/6355 and 6773/6774) showed a distinct genotype and a large evolutionary distance (**Fig. 4**), indicating that the patient was colonized with two different unrelated strains of MSSA. Thus, our findings suggest that three patients retained the same colonizing MSSA during their stay in the ICU and two patients were colonized by a different strain (**Fig. 4 and Table S2**). Of note, isolate pairs 6204/6043 and 6441/6354 were recovered from different patients in hospital B. The pairs shared a high degree of genomic similarity (<20 SNPs), suggesting transmission of MSSA exhibiting the CzIE among patients of the same unit (**Fig. 4 and Table S2**).

**Fig. 4.**
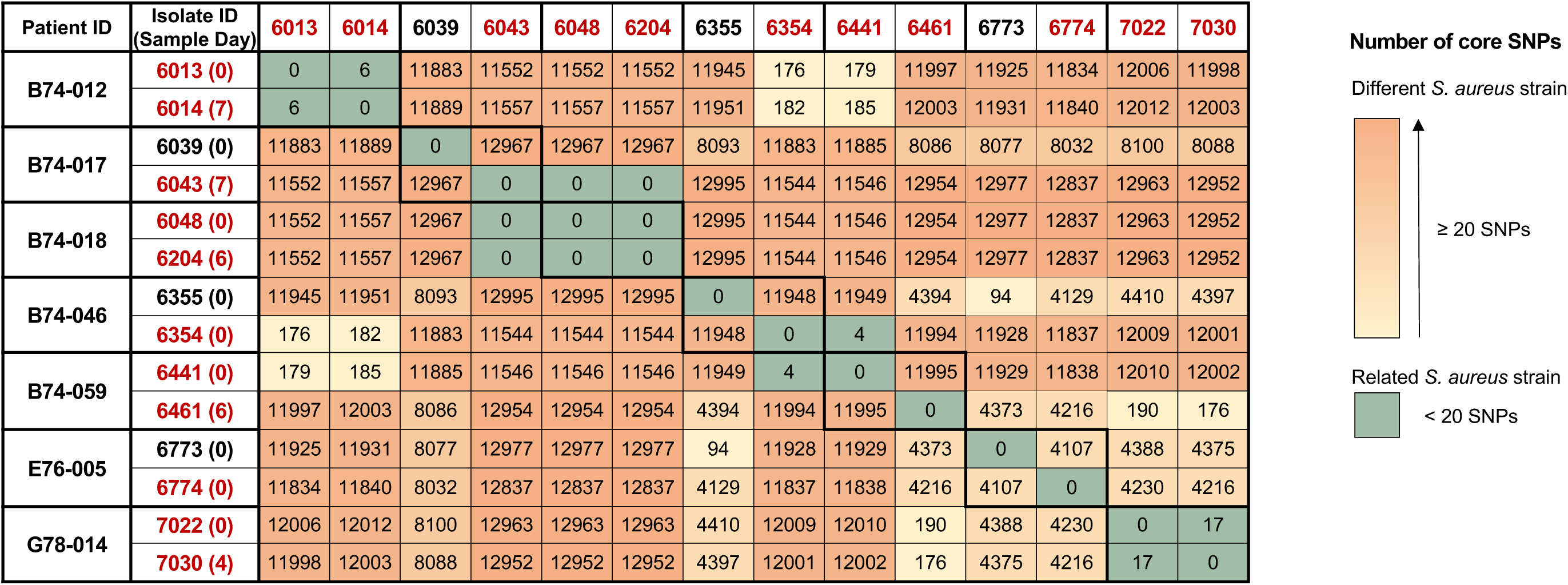
Count matrix of the core SNPs among colonizing MSSA isolates. Core SNPs of the 43 MSSA analyzed (30927 SNPs). MSSA pairs are indicated by white blocks and MSSA isolates with CzIE in red. Cells are colored according to SNPs count between two MSSA: Green, MSSA genomes with < 20 SNPs difference; yellow to orange scale, MSSA genomes with g 20 SNPs likely representing a different strain. Day 0 correspond to admission sampling.

### Components of the nasal microbiota in patients colonized by CzIE positive MSSA had not differences

Using 16S rRNA amplicon sequencing from patients 23 patients (10 colonized by MSSA exhibiting the CzIE and 13 lacking the CzIE), we attempted to identify microbial communities and estimate their diversity and abundance in the context of the CzIE phenotype. At the phylum level, most of the bacterial population (>80%) in both groups consisted of three ASVs (Amplicon Sequence Variants) (*Firmicutes*, *Bacteroidota*, and *Proteobacteria*) (**Fig. S1A**). In both groups, *Firmicutes* was the most abundant taxa followed by Proteobacteria in CzIE-positive group and Bacteroidota in the patients with MSSA lacking the CzIE. No statistically significant differences were observed at the taxonomic level.

At the genus level, we found an abundance of staphylococci (39.4%) and *Bacillus* (3.8%) in the microbiome of patients colonized with MSSA exhibiting the CzIE compared to the patients lacking the CzIE (abundances of 22.9% and 0.04% respectively), Patients colonized with MSSA without the CzIE had a higher abundance of streptococci (3.9 %) and *Moraxella* (3.2%) compared to patients colonized with MSSA showing CzIE (abundances of 1.3 % and 0.06%, respectively) (**Fig. S1A**) No statistically significant differences were observed for these abundances at genus level.

Also, we found no statistically significant differences in α-diversity between MSSA CzIE(+) and MSSA CzIE(-) colonized patients (**Fig. S1B**). However, a lower α- diversity tendency was observed in the MSSA CzIE(+) group, mainly at the genus level. Likewise, the β-diversity estimated using PCoA plots also indicated that the general bacterial population structure was indistinct between patient groups in both taxonomic levels, with a non-significant PERMANOVA (Phylum, p=0.94; Genus, p=0.24) and ANOSIM (Phylum, p=0.96; Genus, p=0.59) analyses (**Fig. S2**). A differential abundance analysis using LefSe showed a unique statistically significant enriched genus (*Anaerococcus* spp.) (p=0.039, LDA score=4.34) in the nasal microbiome of colonized patients with MSSA exhibiting the CzIE (**Fig. S1C**), with a significant major relative abundance in this group of patients.

## DISCUSSION

We postulated that antibiotic pressure, particularly the use of broad-spectrum β- lactam antibiotics may favor the emergence of the CzIE effect in colonizing *S. aureus*. Our findings support our hypothesis, since we found a high point prevalence (58%) of nasal colonization with MSSA exhibiting the CzIE in our multicenter cohort of ICU patients in Colombia. Previous studies^19,22,23^ have shown a high frequency of the CzIE in Colombian MSSA isolates recovered from patients with bacteremia and bone and joint infections and our study confirms that this phenotype seems to be prevalent in Colombian *S. aureus,* although regional variability in the frequency of the CzIE have been documented in MSSA isolates recovered from bacteremia.^19,20,22,23,50–52^ Indeed, among the Latin-American countries (a region with high prevalence of this phenotype) Colombia seems to have one of the highest rates of the CzIE in the region for reasons that are not clear.^22,23^ Data in critically ill and, particularly ICU patients, are scarce and our findings provide some support that this phenotype can be frequently found among critically ill patients that experience heavy antibiotic administration.

The genomic analyses revealed that nasal colonizing MSSA from ICU patients exhibiting the CzIE possessed similar characteristics as previously documented in MSSA isolates with this phenotype and recovered from invasive infections.^21,22^ Moreover, our findings support the notion that nasal colonization in the ICU is a very dynamic process. Indeed, we found that patients in the ICU are commonly colonized with more than one strain of *S. aureus* during the course of the ICU stay. In patients who underwent more than one sampling during the ICU stay, we could determine instances in which the patients were colonized with either the same or different strains. Moreover, we were able to isolate at least two different *S. aureus* strains from the same swab in two patients. Most importantly, our phylogenomic studies provide support of potential transmission of MSSA strains exhibiting the CzIE between different patients in the ICU in the same unit (**Fig. 4**).

Our findings also support a clear association of the CzIE with CC30, harboring BlaZ type A, allotype-2 with Agr type III. We have previously documented this association in isolates from bloodstream infections in Latin America and has also been confirmed in isolates from Korean hospitals.^52–54^ Of note, in a cohort study of pediatric patients with acute osteomyelitis, MSSA exhibiting the CzIE and possessing Agr-III were associated with progression to chronic osteomyelitis.^21^ Of interest, alteration of Agr function has been identified in MSSA displaying the CzIE.^50,54^ Indeed, Agr is responsible of a signaling mechanism in response to population density that plays a preponderant role in activation of expression of virulence factors including the repression of cell-wall associated determinants in *S. aureus*. Furthermore, perturbation in the activity of Agr in *S. aureus* has been associated with decreased activity of several antibiotics (ie. β-lactams, vancomycin).^55^ Mutations in *agr* were found in MRSA isolates exhibiting reduced vancomycin susceptibility phenotypes (ie, hVISA, VISA).^55^ In addition, upregulation of *agr* was identified in *S. aureus* clinical strains under the presence of sub-inhibitory concentrations of oxacillin.^56^ Thus, variations in Agr may be associated with the CzIE and this phenotype could also be marker of enhanced virulence in certain genetic backgrounds.

Although no significant differences were seen in the alpha and beta diversity analyses between CzIE-positive and CzIE-negative nasal microbiomes, *Anaerococcus* spp. were found to be enriched in the CzIE-positive group. Previous studies have described *Anaerococcus* and other genera (*Corynebacterium, Finegoldia, Peptoniphilus, Propionibacterium, and Staphylococcus*) as typical health-associated human upper respiratory tract (URT) bacteria, with a depletion in pathological conditions as chronic rhinosinusitis (CRS).^57,58^ Other studies have shown that anaerobic bacteria, including *Anaerococcus* are more abundant in middle meatuses of nasal cavities in CRS patients compared to healthy subjects.^59^

Thus, the relationship of *Anaerococcus* spp. colonization with MSSA exhibiting the CzIE in the nose needs to be further investigated.

The CzIE is difficult to detect in the routine clinical microbiology laboratory. Indeed, the cefazolin MICs at high inoculum are cumbersome to perform and interpret. We previously developed a rapid nitrocefin colorimetric test based on the observation that we could detect high activities of BlaZ released to the milieu in strains that exhibit this phenotype.^28,29^ In a validation study that included a large collection of Latin American strains, we found that the test had an overall specificity and sensitivity of 88.9% and 82.5%, respectively.^28^ However, we found that ampicillin powder (used to prepare a solution to induce *blaZ*) was not widely available and could be expensive in low resource setting laboratories. Thus, we decided to replace the powder by using an ampicillin disk with a period of incubation of 10 min in BHI broth to release enough ampicillin to serve as the inducer solution. Using the isolates recovered from patients in the ICU, we performed an initial validation of the rapid modified nitrocefin test. Compared with the gold standard (cefazolin MICs at high inoculum), the rapid nitrocefin test exhibited an accuracy of 97.7% (100% sensitivity and 94% specificity). Moreover, the previous version of the test had issues with isolates that harbored Type C BlaZ. The new modified version of the test seems to perform better for type C enzymes with an accuracy of 100%. Of note both type A and C BlaZ enzymes are highly associated with the CzIE.^22,23,28^ The reasons for this discrepancy are not clear and we are currently performing a large validation study to determine specific metrics of the modified test.

Our study has several important limitations, as follows: ***i***) we included only six participant centers in different cities in Colombia that may not reflect the situation and complexity of ICUs in other regions. Nevertheless, the included centers have different levels of complexity and capabilities; ***ii***) the access to clinical information was limited, therefore, the clinical impact of our findings is unknown; ***iii***) we were unable to recover bacterial isolates from patients which developed infections due to logistic difficulties in the clinical laboratories of the participanting centers and, ***iv***) microbiome analyses were only performed in representative samples of patients colonized with MSSA, since we only were able to sequence and analyze 23 samples.

In summary, we found a high point prevalence of the CzIE in colonizing MSSA isolates in ICU patients. Our findings suggest that a complex and dynamic process of colonization of MSSA isolates occurs in ICU patients likely influenced by antibiotic and infection control practices with evidence of patient-to patient transmission of CzIE MSSA strains. The clinical relevance of these findings is the subject of subsequent studies.

## Data Availability

The genomes of the Methicillin-Susceptible Staphylococcus aureus isolates, and the nasal microbiome sequencing were submitted to the NCBI GenBank database under the BioProject numbers PRJNA1107600 and PRJNA1108236, respectively.

## Acknowledgments

The authors thank to Maria Jose Cadavid, Camilo Castellar, Laura Ibarra, Catalina Orejuela and Mauricio Uribe from UGRA, Universidad El Bosque for the technical assistance in this study. Also, to Monica Huertas from Universidad Pedagogica y Tecnologica de Colombia for logistical assistance. This work was supported by Ministerio de Ciencia, Tecnologia e Innovación, grant 130880764152, CT 776-2018 to J.R. and Universidad El Bosque, Convocatoria Interna grant PCI 2023-0002 to J.R. CAA is supported by National Institutes of Health (NIH)/National Institute of Allergy and Infectious Diseases (NIAID) grants K24AI121296, R01AI134637, R01AI148342–01, and P01AI152999.

## Author Contributions

Lina P. Carvajal, Conceptualization, Experimental Assays, Formal analysis, Methodology Coordination, Writing and Original draft; Sandra Rincon, Conceptualization, Experimental Assays, Formal analysis, Methodology Coordination, Writing and Original draft; Sara I. Gomez-Villegas, Methodology Modification and Experiments; Juan M. Matiz-González, Bioinformatic Analysis; Karen Ordoñez, Patient Enrollment; Alejandra Santamaria, Microbiology Testing in Hospital; Leonardo Ospina and Jaime Beltran, Patient Enrollment; Fredy Guevara, Patient Enrollment; Yardany Mendez, Patient Enrollment; Soraya Salcedo, Patient Enrollment; Alexandra Porras and Albert Valencia, Patient Enrollment; Haley Grennia, Alexander Deyanov, Rodrigo Baptista, Sequencing of isolates; Vincent H. Tam, Experimental Assays; Diana Panesso, Experimental Assays and Formal Analysis; Truc T. Tran, Experimental Assays and Formal analysis; William R. Miller, Formal Analysis and Review; Cesar A. Arias, Conceptualization, Writing, Review, Editing and Funding acquisition; Jinnethe Reyes, Conceptualization, Writing, Review, Editing and Funding acquisition.

