## Supplementary material for "Prevalence of the Cefazolin Inoculum Effect (CzIE) in Nasal Colonizing Methicillin-Susceptible *Staphylococcus aureus* in Patients from Intensive Care Units in Colombia and Use of a Modified Rapid Nitrocefin Test for Detection": Figure supplementary 1

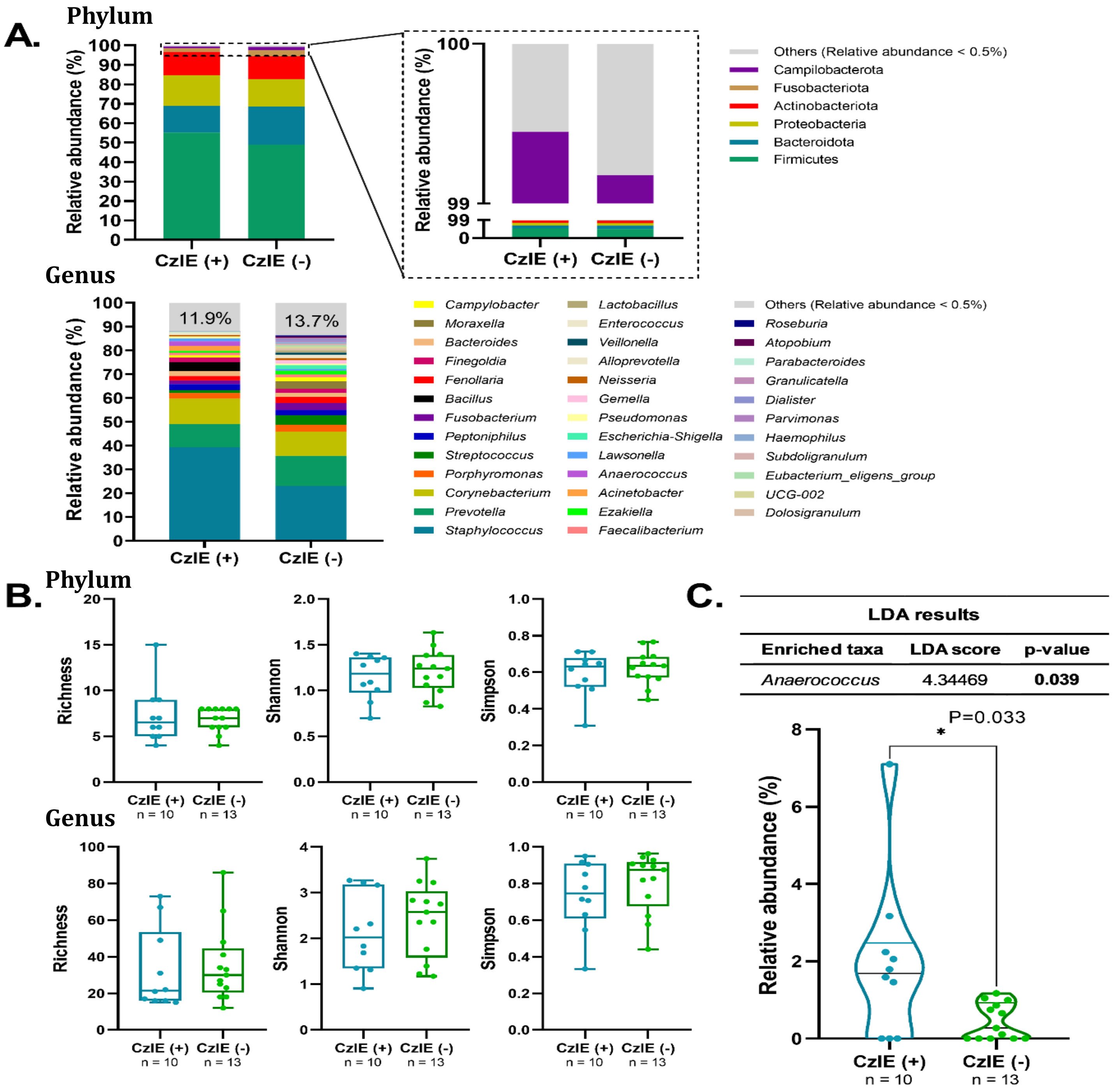
A.
Phylum
Relative abundance (%)
CzIE (+)
CzIE (-)
Others (Relative abundance < 0.5%)
Campilobacterota
Fusobacteriota
Actinobacteriota
Proteobacteria
Bacteroidota
Firmicutes
Genus
11.9%
13.7%
Campylobacter
Moraxella
Bacteroides
Finegoldia
Fenollaria
Bacillus
Fusobacterium
Peptoniphilus
Streptococcus
Porphyromonas
Corynebacterium
Prevotella
Staphylococcus
Lactobacillus
Enterococcus
Veillonella
Alloprevotella
Neisseria
Gemella
Pseudomonas
Escherichia-Shigella
Lawsonella
Anaerococcus
Acinetobacter
Ezakiella
Faecalibacterium
Others (Relative abundance < 0.5%)
Roseburia
Atopobium
Parabacteroides
Granulicatella
Dialister
Parvimonas
Haemophilus
Subdoligranulum
Eubacterium_eligens_group
UCG-002
Dolosigranulum
B.
Phylum
Richness
Shannon
Simpson
CzIE (+) n = 10
CzIE (-) n = 13
Genus
C.
LDA results
Enriched taxa | LDA score | p-value
Anaerococcus | 4.34469 | 0.039
P=0.033
*
Relative abundance (%)
CzIE (+) n = 10
CzIE (-) n = 13

**Fig. S1. Bacterial composition and diversity of CzIE-positive (+) and CzIE-negative (-) MSSA colonized ICU patients**. (**a**) Relative abundance average of the bacterial community at the phylum (top) and genus (bottom) levels. (**b**) α-diversity estimated by the Shannon and Simpson indexes and Richness (observed ASVs) at the phylum (top) and genus (bottom) levels. (c) LEfSe results illustrate microbial taxa enriched in CzIE(+) MSSA colonized patients compared with CzIE(-) group. “Others” category includes all taxa with a relative abundance ≤ 0.5%.
