## Supplementary material for "Prevalence of the Cefazolin Inoculum Effect (CzIE) in Nasal Colonizing Methicillin-Susceptible *Staphylococcus aureus* in Patients from Intensive Care Units in Colombia and Use of a Modified Rapid Nitrocefin Test for Detection": Figure Supplementary 2

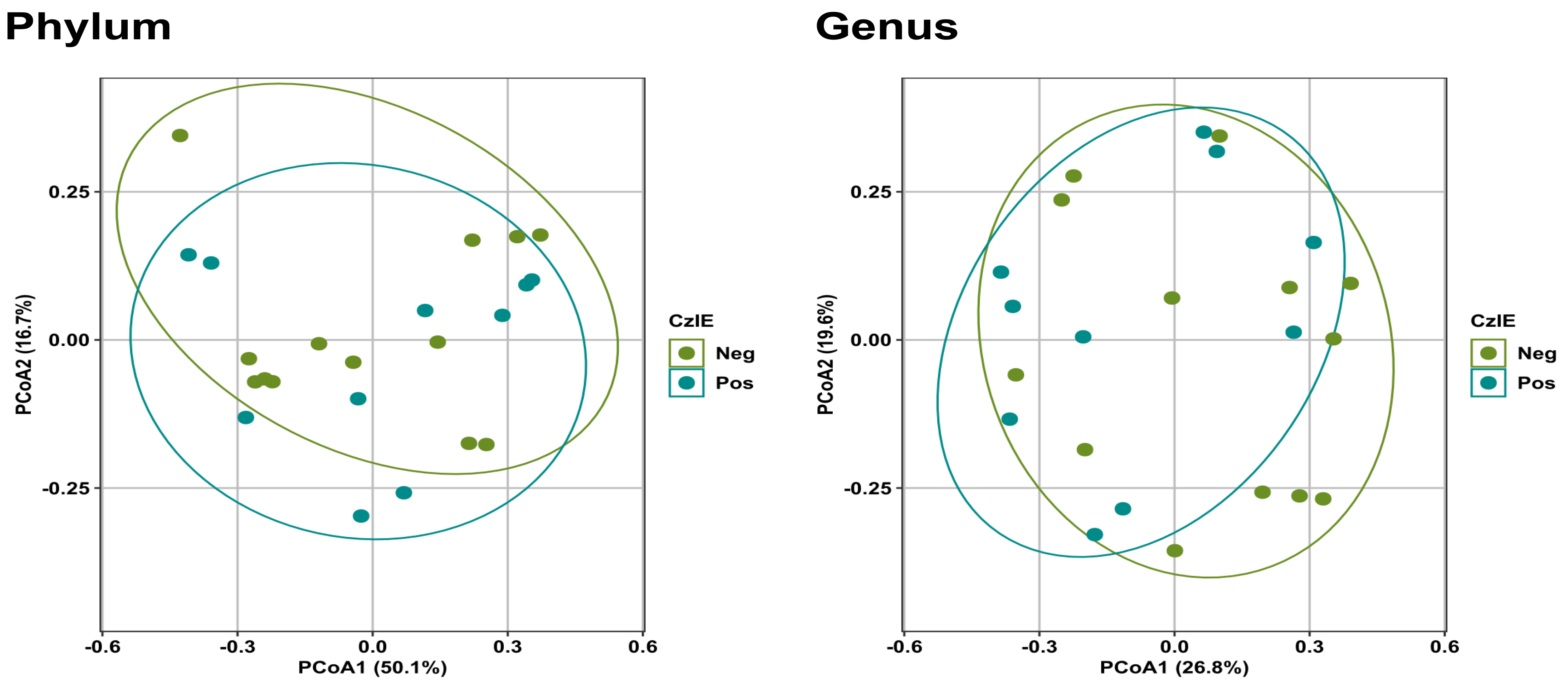


**Fig S2.** Principal Component Analysis (PCoA) of the nasal microbiomes from patient groups colonized with MSSA with the CzIE (**blue**) and lacking the CzIE (**green**) at the phylum and genus levels.
